# On the forecastability of food insecurity

**DOI:** 10.1101/2021.07.09.21260276

**Authors:** Pietro Foini, Michele Tizzoni, Giulia Martini, Daniela Paolotti, Elisa Omodei

## Abstract

Food insecurity, defined as the lack of physical or economic access to safe, nutritious and sufficient food, remains one of the main challenges included in the 2030 Agenda for Sustainable Development. Near real-time data on the food insecurity situation collected by international organizations such as the World Food Programme can be crucial to monitor and forecast time trends of insufficient food consumption levels in countries at risk. Here, using food consumption observations in combination with secondary data on conflict, extreme weather events and economic shocks, we build a forecasting model based on gradient boosted regression trees to create predictions on the evolution of insufficient food consumption trends up to 30 days in to the future in 6 countries (Burkina Faso, Cameroon, Mali, Nigeria, Syria and Yemen). Results show that the number of available historical observations is a key element for the forecasting model performance. Among the 6 countries studied in this work, for those with the longest food insecurity time series, that is Syria and Yemen, the proposed forecasting model allows to forecast the prevalence of people with insufficient food consumption up to 30 days into the future with higher accuracy than a naive approach based on the last measured prevalence only. The framework developed in this work could provide decision makers with a tool to assess how the food insecurity situation will evolve in the near future in countries at risk. Results clearly point to the added value of continuous near real-time data collection at sub-national level.

---

The 2030 Agenda for Sustainable Development, adopted by all United Nations Member States in 2015, calls for urgent action to “end hunger, achieve food security and improved nutrition and promote sustainable agriculture” [1]. However, in 2019, 650 million people were still undernourished [2], with 135 million in 55 countries and territories reported to be acutely food insecure [3]. These numbers have significantly increased as a consequence of the COVID-19 pandemic, with at least 280 million people reported to be acutely food insecure in 2020 [4].

Food insecurity is a complex phenomenon, resulting from the interplay of many environmental, socio-demographic, and political events [5]. It is usually characterized through its four key pillars: availability, access, utilization, and stability; however, agency and sustainability have also been recently proposed as fundamental dimensions of food insecurity [6].

Climate variability and extremes weather events are considered among the main drivers, mostly because of their effects on crop production [7, 8]. In recent years, studies have additionally shown that, beyond agricultural yields, extreme temperatures and precipitation conditions also directly negatively affect child nutrition and food security [9, 10, 11, 12].

Conflicts are also deeply intertwined with food insecurity, of which they can be the cause or the consequence [13]. Several studies have shown that conflict impacts agricultural production, nutritional status, coping and consumption [14, 15].

Food insecurity is also a global public health challenge. Household food insecurity is the leading risk factor of malnutrition, claiming approximately 300 000 deaths each year. Whether directly or indirectly, due to inadequate food consumption and poor diet quality, it is also accountable for over half of all deaths among children in Sub-Saharan Africa. Among the various determinants of food insecurity in relation to child malnutrition, the main ones are socio-demographic characteristics as well as food prices [16]. The impact of food insecurity on health cannot be overstated [17, 18, 19]. The current COVID-19 crisis has exacerbated the situation and undermined some of the critical elements influencing food insecurity, e.g. international agricultural supply chains and goods prices impacted by trade restrictions [20]. In low and middle income countries, social distancing, workplace closures, and restrictions on mobility and trade had cascading effects on economic activity, food prices, and employment [21].

From a policy perspective, food insecurity will remain a worldwide concern for the next 50 years and beyond. As a 20-year old article in Science stated: “Recently, crop yield has fallen in many areas because of declining investments in research and infrastructure, as well as increasing water scarcity. Climate change and HIV/AIDS are also crucial factors affecting food security in many regions. Although agroecological approaches offer some promise for improving yields, food security in developing countries could be substantially improved by increased investment and policy reforms” [22]. In the past twenty years, the situation has however significantly worsened and agroecological technologies are not keeping up with the accelerating effects of climate change that, ever increasingly, are affecting countries and populations on much shorter scales (weeks and months rather than years) than in the past.

Therefore, an essential step towards achieving hunger reduction is to have access to frequent, up-to-date information on the status of food insecurity in countries facing humanitarian crises, and to estimates of where and when the situation is likely to improve or deteriorate, in order to allow for informed and timely decision-making on resource allocation and on relevant policies and programmes. For this reason, governments and international organizations perform food security assessments on a regular basis, by means of face-to-face surveys and, increasingly, through computer-assisted telephone interviews (CATI), or technologies like interactive voice response and web surveys. The World Food Programme (WFP) is currently monitoring the food security situation in near-real time in a number of countries at the sub-national level, collecting on a daily basis, through remote phone surveys, information on levels of food consumption and food-based coping, as well as other relevant indicators [23, 24]. This unprecedented availability of daily sub-national level data paves the way for new possibilities since it not only allows for a continuous up-to-date picture of the current situation, but could also be used to build predictive models to forecast how the situation will evolve in the future. In this study, we explore the forecastability of insufficient food consumption levels, and show, specifically for Syria and Yemen, that satisfactory predictions up to 30 days into the future can be obtained when enough daily sub-national level historical data is available. Having access to regular real-time estimation of how the situation is likely to evolve in the near future would allow WFP for more informed discussions on need-based humanitarian assistance allocation decisions.

Forecasting modeling has been the subject of extensive investigation during the last decade in different fields, from financial markets [25, 26] to infectious disease epidemiology [27, 28, 29, 30]. However, it is still a relatively new area of research in the context of food security. The Food and Agriculture Organization of the United Nations (FAO) developed a methodology to produce annual country-level estimates of the prevalence of undernourishment, and to project these estimates up to 10 years into the future [31, 32]. Okori and collaborators first proposed to use machine learning models to predict whether a household is in famine or not from household socioeconomic and agricultural production characteristics [33, 34]. The effort of predicting levels of insufficient food consumption has been tackled in the context of Malawi, in a study where the authors built a model trained on 2011 data to estimate the situation in 2013 [35], and more recently in a work proposing a model to nowcast sub-national levels of insufficient food consumption on a global scale [36]. Both studies propose methods to predict the current situation when primary data is not available, but they do not address the challenge of making projections for the future. The World Bank recently proposed a machine learning approach to forecast transitions into critical states of food insecurity [37] and a stochastic model to forecast famine risk [38]. Concurrently, similar work was carried out by other researchers in the context of Ethiopia [39]. These studies focus on forecasting month-to-month transitions to different phases of food insecurity, based on the Integrated Food Security Phase Classification (IPC) framework [40].

In this study, we tackle a different problem: forecasting the daily evolution of the prevalence of people with insufficient food consumption at the sub-national level. This metric, characterizing a given area at a given time, is defined as the prevalence of households, in the specified area and time period, that are identified to have poor or borderline food consumption. Such prevalence is computed from a representative number of household surveys enquiring about one of the core household food insecurity indicators, namely the Food Consumption Score (FCS), which captures households’ dietary diversity and nutrient intake [41]. Having access to reliable predictions of the evolution of insufficient food consumption levels over future weeks and months could allow governments and organizations to identify which areas should be monitored more closely and to eventually take timely decisions on resource allocation.

The main difference with IPC and the Famine Early Warning Systems Network (FEWS NET) [42] is that, while these make use of consensus-based expert opinion, our approach is algorithmic and data-driven, and can hence be applied in an automatic fashion. IPC’s and FEWS NET’s projections are essential for humanitarian action, however they require local expertise and considerable time to be developed. Our main goal is not to replace these efforts, but rather to complement them with an approach that, once improved as more data becomes available, could be used to provide rapidly available forecasts for several places at the time by automatically feeding near real-time data to the proposed algorithms.

## Results

### Time trends of insufficient food consumption

We study the possibility of forecasting one of the core dimensions of food insecurity by means of a unique data set of daily sub-national time series of the prevalence of people with insufficient food consumption, in six countries in West Africa and the Middle East: Burkina Faso, Cameroon, Mali, Nigeria, Syria and Yemen (see the Methods for a detailed definition of the indicator under investigation). These countries, although having varying socioeconomic and geopolitical characteristics, have all been identified as major food crises where acute food insecurity is driven by conflict, weather extremes and economic shocks [3]. Among all major food crises, these are the countries for which the largest volume of food insecurity data is available.

The length of these time series varies from a minimum of 865 days in Mali 1340 days in Yemen, over the years 2018-2022. Also, geographic coverage varies across countries. In the case of Burkina Faso, the prevalence of insufficient food consumption is available for all administrative units of the country, while only 3 states of Nigeria are included in our dataset (Adamawa, Borno, Yobe), given these are the most at risk areas closely monitored by WFP. Overall, our dataset covers 88% or more of the total population in all countries, with the only exception of Nigeria (see Supplementary Table S4).

Since the mode of questionnaire administration can have serious effects on data quality [43, 44], during the last few years WFP conducted mode experiments in several countries, each demonstrating the feasibility of collecting food security indicators via CATI surveys [45, 46]. However, sampling and selection bias should be assessed and mitigated [47, 48, 49], hence post-stratification weights were applied by WFP, as detailed in the Methods section.

As shown in Fig. 1, in the six countries, time trends of insufficient food consumption display noisy and irregular patterns, underscoring the complex dynamics underlying food insecurity. During the study period, all countries experienced large fluctuations in the prevalence of insufficient food consumption, and such variations were not uniform between sub-national administrative units. In Cameroon, for instance, only a few regions were characterized by a relatively high proportion of food insecure people, generally above 50%, but also exhibiting large fluctuations, such as the rapid decline and subsequent increase observed in the North-West regions. On the other hand, in Syria, the sub-national trends were all similar in terms of relative changes in the affected population, with a general upward trend affecting almost every province beginning in July 2020. In the governorates of Yemen, for which the longest time series are available, the proportion of the population affected by food insecurity varied between 20% and 60% during the years 2018-2022, however, a common national time trend is less recognizable. It should be noted that some of these irregularities could also be partially due to the effects of sampling and selection bias, whose mitigation can rarely be achieved in full.

**Figure 1:**
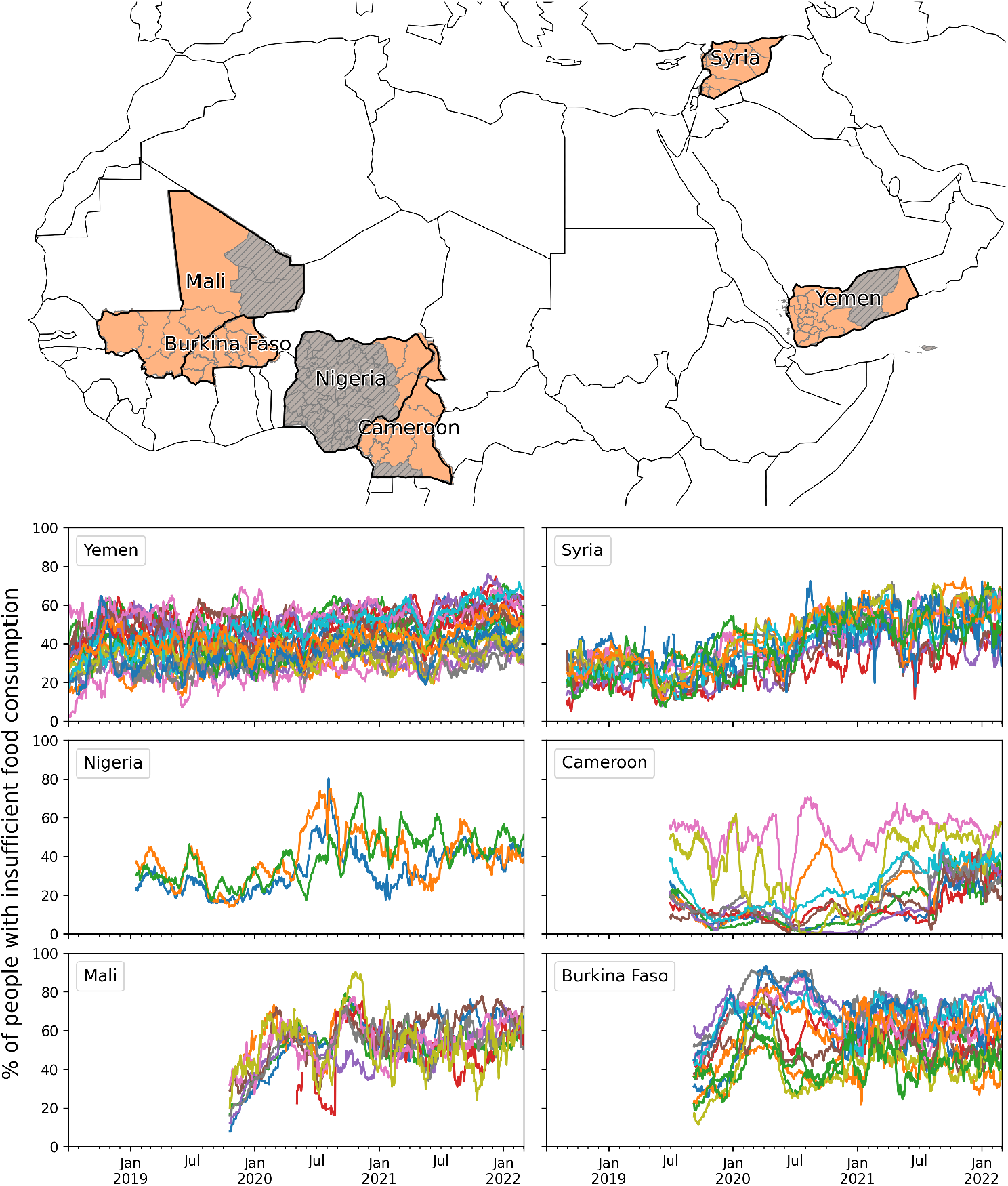
Time trends of insufficient food consumption. Each panel displays daily time series of the percentage of people with insufficient food consumption in the first-level administrative units of Burkina Faso, Cameroon, Mali, Nigeria, Syria and Yemen. The six countries are highlighted in the map, and the orange shade indicates the areas that are considered by our analysis.

### Permutation entropy and intrinsic predictability of food insecurity

We first quantify the intrinsic predictability of the time series shown in Fig. 1 by means of a permutation entropy analysis. Permutation entropy (PE) is a model-free measure of time series complexity [50, 51], that is conceptually similar to the Shannon entropy but is based on the frequency distribution of motifs. PE has been extensively used to assess the predictability of time series in different domains including finance and economics [52, 53], ecology [54] and infectious disease epidemiology [30]. In short, to compute the PE of a time series we translate its real valued sequence (*x*_1_, *x*_2_, …, *x*_*N*_) into a frequency distribution of symbols that represent patterns of relations *x*_*i*_ *< x*_*j*_, *x*_*i*_ = *x*_*j*_ or *x*_*i*_ *> x*_*j*_ between nearest or distant neighbors, *x*_*i*_ and *x*_*j*_. Such frequency distribution is then used to assess the predictability of the time series by computing the Shannon entropy associated with the distribution of permutation patterns in the symbols defined above. In the Methods section we provide a complete formal definition of the PE and its computation. It has been shown that PE can be considered as a measure of intrinsic predictability of a time series and its value is positively associated with forecasting error [54]. Intuitively, PE quantifies the information that is transmitted from the past to the present state of a time series: a time series that periodically visits the same few symbols among the many possible will have a low entropy and its present state will be easily determined from the past. A random time series that uniformly samples the symbols with equal probability will have a high entropy and its future will not be predictable from past states.

In the case of food insecurity, we find that insufficient food consumption trends are not easily predictable based on their past history. As shown in Fig. 2, their predictability, measured as *χ* = 1 − *H*, where *H* is the PE, never reaches values above 0.5 and it is often reduced to 0.1 − 0.2 within a 10-day horizon. These values are generally much lower than those observed in the case of infectious disease dynamics [30] and they are closer to measures of predictability of financial time series [55], which are characterized by a high short- and long-term volatility. Confidence intervals around mean predictability values are also narrow, highlighting a consistent lack of recurrent patterns in the insufficient food consumption time series across different time scales, which in turn highlights the presence of intrinsic entropy barriers to their predictability.

**Figure 2:**
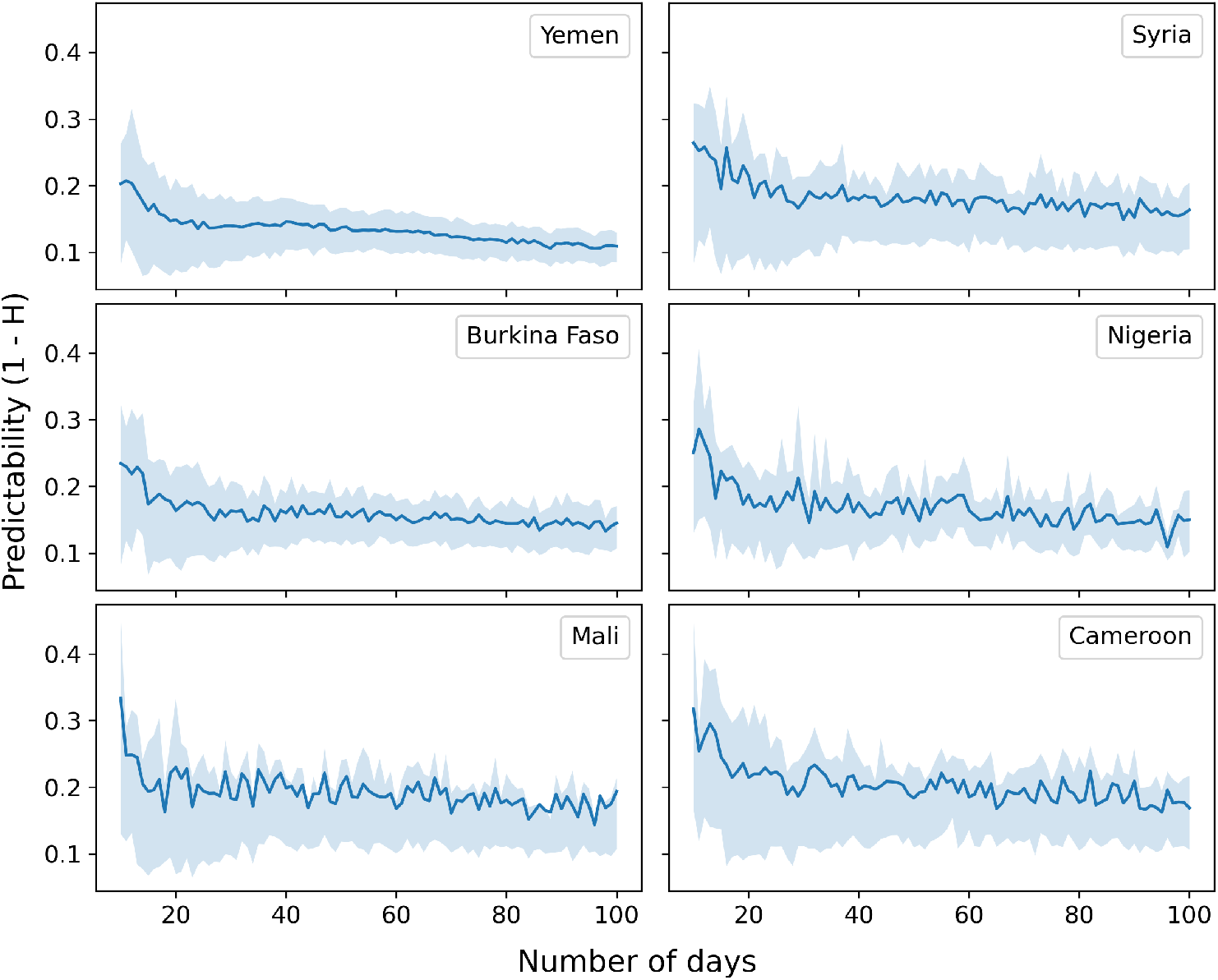
Food insecurity is characterized by low predictability. The average predictability *χ* = 1 − *H* for daily trends of insufficient food consumption, in the six analyzed countries, is shown as a function of time series length in days. We average *H* over temporal windows by selecting 1,000 random points from each time series and calculating *H* for windows of length 10, 11, 12, …, 100 days. The solid lines indicate the mean value and the shaded areas mark the interquartile range across all administrative units and starting locations in the time series.

### Forecasting food insecurity with secondary information

Following from the observation that insufficient food consumption trends are not highly predictable from their own history, we explore whether secondary information can be used to enhance our ability to predict their future evolution. To this end, we revert to information on the key drivers of food insecurity: conflict/physical insecurity, extreme weather events and economic shocks [56]. We build a set of indicators covering these three domains and develop a forecasting model based on gradient boosted regression trees (XGBoost) [57] to make predictions on how the insufficient food consumption trend will evolve up to 30 days into the future. More specifically, in our model we consider as predictors of insufficient food consumption the following indicators (see Methods and the Supplementary Information file for a full description). First, we include daily time series of the prevalence of people using crisis or above crisis food-based coping, which is obtained from another core food insecurity indicator, the reduced Coping Strategy Index (rCSI), by measuring the share of households with rCSI ≥ 19 [58, 40]. Since political unrest can affect food security, we include in our model daily time series of fatalities due to conflict or political violence as reported by the Armed Conflict Location and Event Data Project (ACLED) [59]. Economic shocks are included into the model by considering monthly variations in the price of cereals and tubers in local currency. The model takes into account the effects of weather events and climate conditions by including time series of rainfall, of its anomaly with respect to long-term averages (over 1 and 3 months), and time series of the Normalized Difference Vegetation Index (NDVI), a standard satellite-based measure of vegetation coverage that is commonly used for drought assessment [60], and of NDVI anomaly. Finally, since the food consumption behavior of most of the population in several African and Asian countries is affected by Ramadan, we include a time series that marks the days of the Ramadan period that fall within the time window used to measure people’s food consumption.

Fig. 3 and Supplementary Fig. S1 show the prediction results of the model for the case of Yemen and Syria, respectively, the countries for which the longest time series of insufficient food consumption prevalence are available. In Yemen (Syria), cross-validated predictions can explain between 99% (99%) and 72% (47%) of the variation in insufficient food consumption, with the former being the variation explained by the 1-day into the future forecast, and the latter for the 30-day into the future one (Fig. 3a and S1a). This is a significant increase of *R*^2^ with respect to a naive prediction based on the last measured value only, which can only explain between 99% (99%) and 65% (31%) of the variation (Fig. 3c and S1c), and whose mean squared error (MSE) is larger and with a wider dispersion than the MSE of the proposed model (Fig. 3b,d and S1b,d). The scatterplots in Fig. 3e and S1e show the performance of the forecasting models as the predicted insufficient food consumption value against the actual value, for different prediction horizons. As expected, dots get further away from the identity diagonal as the prediction horizon increases up to 30 days, although the general behavior is consistent with a good predictive accuracy.

**Figure 3:**
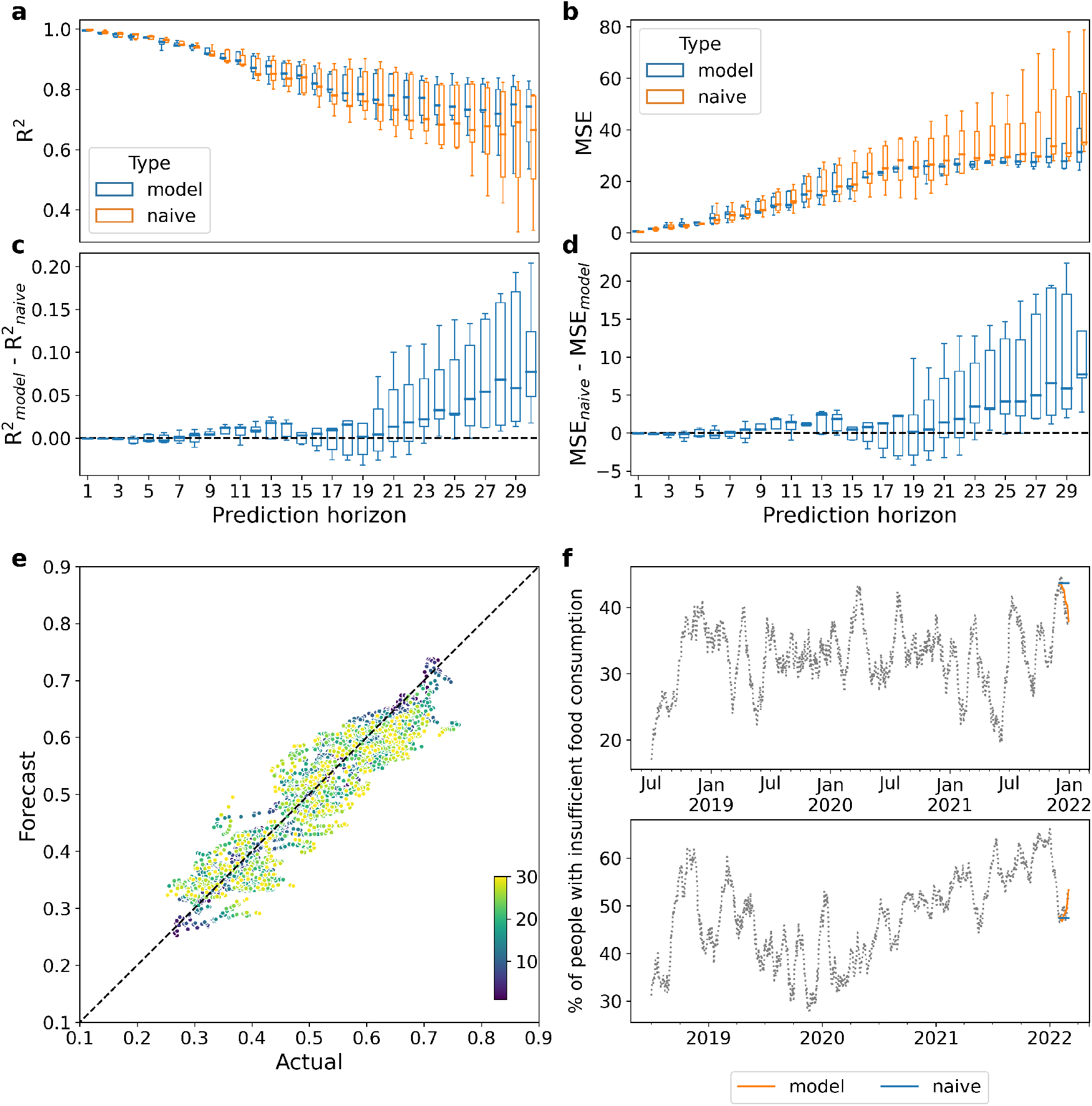
Forecasting the prevalence of people with insufficient food consumption in Yemen. The forecasting is performed over 5 different monthly splits of all governorates time series, from October 2021 to February 2022. (a) Box plots of the coefficient of determinations (*R*^2^) across the 5 splits for both the proposed and the naive models (in blue and orange, respectively), for each forecasting horizon. (b) Box plots of the mean squared error (MSE) across the 5 splits for both the proposed and the naive models for each forecasting horizon. (c) Box plots of the difference between the *R*^2^ of the proposed and of the naive model for each split. (d) Box plots of the difference between the MSE of the naive and of the proposed model for each split. (e) Predicted vs actual value for each data point in the 5 splits. Colors represent the corresponding forecasting horizon and vary from dark blue (1 day) to yellow (30 days) (f) Example of forecasting results for December 2021 in Amanat Al Asimah (top) and February 2022 in Abyan (bottom).

Over short forecasting horizons, typically less than 14 days, a naive approach proves to be a good enough predictor as we do not expect food consumption to suddenly change from one day to the next. However, as we try to forecast further into the future, we see that the forecasting model starts to outperform the naive approach, as shown in Fig. 3f (S1f) for the case of two Yemeni (Syrian) provinces over a 30 day horizon.

In the case of the remaining four countries, whose available training points are less than half than for Yemen, the model performance is worse than the naive approach across all prediction horizons, both in terms of *R*^2^ and *MSE* (see Supplementary Figs. S2 - S5).

### Model performance as a function of data availability

Given the relatively poor predictive performance of our models in countries with short time series of insufficient food consumption, we systematically examine how the performance varies as a function of the length of the time series available to train the model and of spatial coverage, indicating the number of sub-national areas. We find that, compared to the naive approach, the performance of our model dramatically increases with the number of available training points, which is given by the product of the two dimensions above: temporal length and spatial coverage (see Fig. 4). Moreover, with a given size of the training set, the proposed model tends to perform better than the naive approach as the forecasting horizon grows, demonstrating that, as expected, the model is better at predicting further into the future than just considering the last available measurement. However, this effect is evident only when a large training set is available (as in the case of Yemen, with more than 20,000 data points), and a small training set reduces the benefit of the model even over longer time horizons.

**Figure 4:**
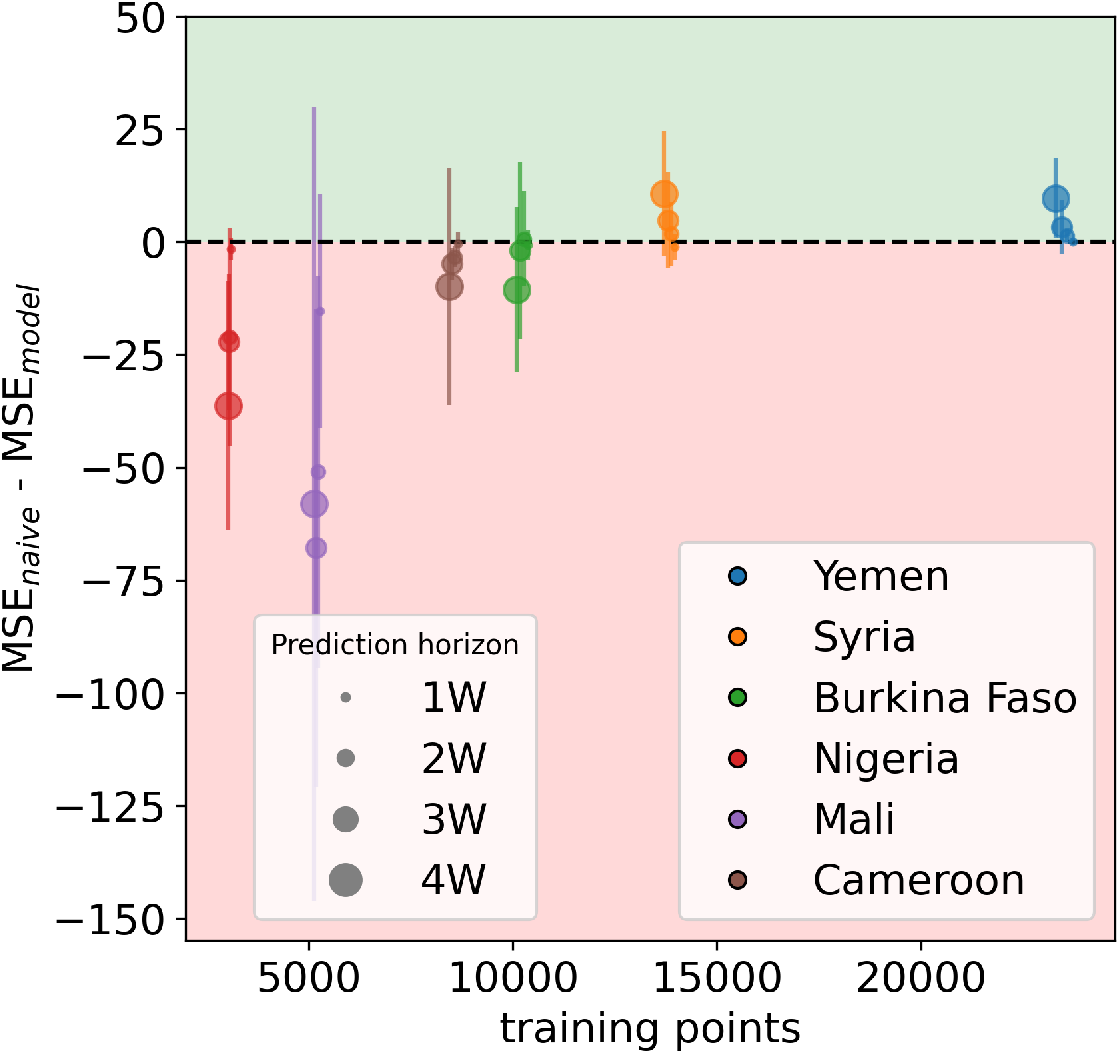
Model performance as a function of the number of available training points. For the six analyzed countries and four different forecasting horizons (1 to 4 weeks), the figure shows the averaged differences between the MSE of the naive approach and the MSE of the forecasting model across the different splits, as a function of the size of the training set. Error bars correspond to the relative standard deviation. The green area indicates where our model outperforms the naive one, the red area indicates the opposite.

A limitation of this analysis relies on the fact that in our dataset different numbers of training points coincide with different countries, which does not allow to disentangle effects due to the local context from those due to temporal length and spatial coverage. We therefore performed further analyses in order to separately investigate the role played by the number of covered areas and by the time series length. First, we considered the same time series length for all countries by using as starting date for all of them the earliest date available for all countries. In this setting, the difference in the number of training points among the different countries is only due to the different number of areas covered. Results are reported in Supplementary Fig. S6a. Secondly, we considered instead the full time series but we fixed the number of considered areas to the minimum number of areas available for all countries (in this case we had to exclude Nigeria since only three areas are covered there, which would have been too few for the analysis). In this setting, the difference in the number of training points among the different countries is only due to different lengths of the available time series. Results are reported in Fig. S6b. In both cases, we re-trained and tested the model using, each time, a different data subset, as described above. Results show that the performance of the model still increases with the dimension under study (the length of the time series and the number of covered areas), confirming our initial results.

## Discussion

In this study we addressed the critical challenge of forecasting the daily evolution of a food security indicator, namely the prevalence of people with insufficient food consumption, as measured by WFP. The problem promptly proved difficult given that the analyzed time series exhibit noisy and irregular behavior. This is to be expected since food insecurity in Sub-Saharan Africa and the Middle East is a highly dynamic phenomenon, comprising a seasonal component related to agricultural production calendars and religious observances such as Ramadan (during which consumption patterns are completely altered), but also subject to swift changes when external shocks hit, such as the emergence of conflict, extreme weather events or economic shocks [61, 62, 63].

Therefore, forecasting based solely on information on the historical evolution of the target indicator over time would not be successful, as demonstrated through a permutation entropy analysis. Hence, we extended the proposed framework to include historical information on the key drivers of food insecurity and built models that comprise both endogenous (insufficient food consumption itself, as well as food-based coping information) and exogenous factors (conflict-related fatalities, rainfall and vegetation and their anomalies, staple food prices and Ramadan’s occurrence). We showed that the proposed model makes it possible to forecast the prevalence of people with insufficient food consumption up to 30 days into the future with higher accuracy than a naive approach solely based on the last measured prevalence, at least in places where enough training data are available to inform the model. The number of available historical observations proved to be a key element in forecasting success. Even for places with more than 10,000 available training points, which is not an extremely large number but still enough to provide reasonable results in other contexts [64], the phenomenon seems to be too complex for the algorithms to learn meaningful patterns.

Besides the external shocks related to local socio-economic conditions, it is important to note that all countries under study experienced the global effects of the COVID-19 pandemic since early 2020. The pandemic has significantly impacted food security on a global scale [65], disrupting supply chains, limiting access to food due to the adoption of non-pharmaceutical interventions, and increasing the need for food assistance [66]. In our analysis, we did not include epidemiological variables to model the impact of the COVID-19 pandemic – such as reported cases or deaths – because those epidemiological indicators are likely to be unreliable in the countries under study, due to the lack of adequate surveillance systems [67]. On the other hand, we expect the effects of the pandemic to be mainly captured by market price trends [68], which indeed markedly increased in all countries and in all regions as shown in Fig. S9 of the Supplementary Information. As SARS-CoV-2 continues spreading worldwide, the world economy still suffers from the consequences of the pandemic and its long term effects are hard to predict. Further research will be needed to investigate the complex interplay between the COVID-19 pandemic and food security.

Forecasting research within the humanitarian context has only recently started to attract attention from scholars [69]. In this context, our study represents an initial step towards the application of forecasting approaches to food insecurity at a high spatial and temporal granularity. Our results confirm that nowcasting or one-step-ahead forecasting are feasible, as reported in recent studies [36, 38], but long-term forecasts are challenging and strongly conditioned by data availability. The methods presented in this study come with limitations, and they could be further improved through several approaches. First, more complex forecasting methods could potentially lead to a greater forecasting accuracy, for instance through the use of deep learning techniques [70]. Additionally, hybrid methods, combining both statistical and ML features, could achieve a better forecasting performance [64]. Finally, forecasting models could benefit from the inclusion of additional external predictors and in particular from the availability of novel data streams, such as mobile phone data or the automated text mining of news [72].

In conclusion, our study presents a simple, yet fundamental message for governments and humanitarian organizations on the power of the data they collect: collecting data on a regular basis for long enough periods of time and across enough different geographic areas does not only make it possible to monitor the evolution of a situation in near real-time but also to inform forecasting models that would make it possible to produce estimates of how the situation is likely to evolve in the near future. This means that decision makers would have access in advance to information on areas most at risk of a deterioration in the food security situation, allowing for a more timely response. Predictions should of course be used with caution and considered only as an indication of what may happen in the near future, hence informing preparedness efforts by suggesting a need for further in-depth assessments of the food security situation.

## Materials and Methods

### Target indicator

The indicator whose time-evolution we aim to predict is the daily prevalence of people with insufficient food consumption in a given sub-national geographical area. This prevalence is obtained as the weighted share of households in the area that are found to have poor or borderline food consumption according to the Food Consumption Score (FCS) [41, 73]. The FCS is obtained through household surveys by asking how often, during the previous 7 days, a household has consumed food items from different food groups (main staples, pulses, vegetables, fruit, meat and fish, milk, sugar, oil and condiments). Consumption frequencies are then summed up in a weighted fashion, where each food group is weighted according to its nutritional level (with more nutritious foods having higher weights), resulting in the FCS. Thresholds are then applied to label each household as having poor, borderline or acceptable food consumption (as further detailed in Section A of the Supplementary Information), allowing to eventually compute the prevalence of people in a given area with insufficient (i.e. poor or borderline) food consumption.

The time series analyzed in this study are obtained through daily computer-assisted telephone interviewing (CATI) surveys. Sample sizes were determined by WFP by taking into account modality and adhering to IPC guidelines for a good level of reliability (i.e. as close to 150 households per strata as possible) [40]. They initially utilize Random-Digit Dialing (RDD) to obtain the most random selection of respondents as possible, while applying some filters to ensure the required geographic and sociodemographic distributions (i.e. not all households reached are actually interviewed, only those matching the specific characteristics needed). This enables WFP, over time, to build a representative sample, and then transition to panel surveys after the initial months of implementation [46].

In order to compute a statistically representative prevalence of people with insufficient food consumption at sub-national and daily resolution, a rolling window approach is used. That is, for each geographical area, the prevalence of people with insufficient food consumption for a given day is obtained as the weighted share of households with poor or borderline food consumption interviewed during the previous *d* days, where *d* varies by country (values are reported in Supplementary Table S2). Missing values in the time series are inferred through linear interpolation. Post-stratification weights are applied by WFP to compute the final share of households with poor or borderline food consumption in a given area and time window, in order to mitigate sampling and modality bias, as detailed in [46]. This is done through weighting of the data to account for the under-representation of certain demographics. Population weights are applied to compensate for administrative areas that are under- or over-sampled, while demographic weights are introduced to mitigate selection bias and compensate for under-represented households (e.g. low-income or less-educated households). The final weight is given by the product of the two, when both need to be applied.

### Permutation entropy

We employ the *Permutation Entropy* (PE) as a model-free measure of time series predictability [30]. The main assumption of this approach is to measure the Shannon entropy through the probabilities of encountering trend patterns within the time series. For this reason, the PE first categorizes the continuous time series *X* in a small set of symbols or alphabet according to their trends. Let *x*(*i*), *i* = 1, …, *N*, denote sequences of observations from a system *X*. For a given, but otherwise arbitrary *i, m* amplitude values *X*_*i*_ = {*x*(*i*), *x*(*i*+*τ*), …, *x*(*i*+(*m* − 1)*τ*) } are arranged in an ascending order where *τ* denotes the time delay, and *m* is the embedding dimension. Each *X*_*i*_ is then mapped onto one of the *m*! possible permutations. The PE of the time series *X* is given by the Shannon entropy on the permutation orders:

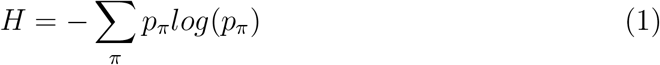

where *p*_*π*_ is the probability of encountering the pattern associated with permutation *π*. An important convenience of symbolic approaches is that they discount the relative magnitude of the time series [74]. This is important in our case because different geographical units can differ largely in food insecurity prevalence. The embedding dimension *m* and the time delay *τ* are to be set in order to derive a reliable state space. There exist different procedural approaches in order to deal with this setting decision [75, 76]. In order to find the appropriate embedding dimension for clustering a set of time series, we follow the instructions proposed by Scarpino & Petri [30]. The time delay is fixed to *τ* = 1 in order to get results from continuous intervals. Finally, the metric used is the predictability defined as *χ* = 1 − *H*. The closer to 1 the *χ* is, the more regular and more deterministic the time series is. Contrarily, the smaller *χ* is, the more noisy the time series is. As suggested by Scarpino & Petri [30], we analyzed the predictability as a function of the length of each time series. Focusing on the predictability over short timescales, we average *H* over temporal windows by selecting 1,000 random points from each time series and calculating *H* for windows of length 10, 11, 12, …, 100 days.

### Independent variables

The following variables were defined to be considered as input features for the forecasting models.

#### Prevalence of people using crisis or above crisis food-based coping

This prevalence is obtained as the weighted share of households in a given sub-national geographical area that are found to have a reduced Coping Strategy Index (rCSI) greater than or equal to 19 [58, 73]. The rCSI is obtained through household surveys by asking if and how often, during the previous 7 days, a household had to adopt the following coping behaviors: relying on less preferred or less expensive food, borrowing food from relatives or friends, limiting portion sizes, restricting adults’ consumption in order for small children to eat and reducing the number of meals eaten in a day. The rCSI is then obtained as a weighted sum of these frequencies, where weights are based on the severity of the strategy, as further detailed in Section B of the Supplementary Information. The survey data used to build this variable is the same as for the target indicator. A rolling window approach to compute a statistically representative prevalence of people using crisis or above crisis food-based coping at sub-national and daily resolution is also applied, and missing values are interpolated through linear regression. The same post-stratification weighting schemes to mitigate sampling and modality bias are also applied.

#### Conflict-related fatalities

The number of conflict-related fatalities in a given geographical area is obtained from the Armed Conflict Location and Event Data Project (ACLED), a publicly available near-global repository of reported conflict events and related fatalities [59]. Since each daily value of the target indicator is based on data collected during the previous *d* days, the number of fatalities associated with the same date and area is also obtained by summing all fatalities reported in the same area during the same *d* days. Further details are reported in Section C of the Supplementary Information.

#### Market prices

Monthly prices of cereals and tubers are obtained from WFP’s publicly available Economic Explorer^*^. Cereal and tubers prices for each geographical area and date are obtained by averaging normalized prices (in local currency) across all markets within the area. Further details are reported in Section D of the Supplementary Information.

#### Weather variables

In order to measure the performance of the agricultural season, and more specifically whether the rainfall season is drier or wetter than average, and its impact on the vegetation status, for each geographical area and date, we consider the following weather variables, which are defined and computed by WFP as 10-day measurements, for each first-level administrative unit, and made publicly available through its Seasonal Explorer^†^: the amount of rainfall in mm, its 1-month and 3-month anomalies with respect to the historical average during the same period of the year (expressed in percentage), the normalized difference vegetation index (NDVI), and its anomaly (defined as for rainfall but considering 10-days only since effects of previous rainfall are already integrated by vegetation itself). Further details are reported in Section E of the Supplementary Information.

#### Religious observances

Ramadan is a religious observance celebrated by that the majority of the population in the analyzed countries during which food consumption increases. For each date and geographical area we therefore create a variable that takes into account the number of days, within the *d* days considered to obtain the prevalence of people with insufficient food consumption for the same date and area, that fall within the Ramadan observance period. This variable therefore spans between 1 and *n* during and after Ramadan, and is otherwise equal to zero during the rest of the year.

#### Population

The latest population estimate provided by WFP for each geographical area is used as a static variable.

#### Geographical area identifiers

The total area of each geographical unit, its latitude and longitude, and it waterways size, are also used as static variables.

#### Temporal identifiers

Temporal information (day, month and year) on the forecasting horizon is also included.

### Preliminary feature selection

A correlation analysis based on Pearson’s *ρ* and on the Variance Inflation Factor (VIF) was performed on this initial set of variables for each individual country in order to detect highly collinear features, as further detailed in Section 5 of the Supplementary Information. As a result of this process, the rainfall 3-month anomaly was removed from all country-specific datasets but Nigeria’s, the NDVI anomaly from Yemen’s and Syria’s, and the NDVI from the remaining four countries (see Supplementary Table S6), leaving rainfall, the 1-month rainfall anomaly and NDVI or its anomaly as the only remaining weather-related features for most countries, given the low level of correlation between them.

### Forecasting

The core of this work revolves around the forecasting effort focusing on predicting the evolution up to 30 days into the future of our insufficient food consumption time series^‡^. To this aim, we chose to use the *eXtreme Gradient Boosting* (XGBoost)^§^ algorithm [77], a widely used state-of-the-art machine learning technique known for its high performance and flexibility. XGBoost belongs to the category of so-called ensemble learning approaches, which is a branch of machine learning methods that makes use of several models at once to produce a single better output. In the case of XGBoost, the base model is a decision tree, which is considered best-in-class for handling small to medium-sized data. A decision tree is a set of hierarchical choices which eventually lead to a final result, i.e the prediction. Ensemble methods combine several decision trees to produce better predictive performance than utilizing a single decision tree. To create this collection of trees, XGBoost fits consecutive trees by, at every step, trying to solve for errors from the previous tree, using a gradient descent algorithm. The wider context of machine learning approaches used in the time series forecasting field and a more in-depth description of XGBoost can be found in Section 6 of the Supplementary Information.

The motivation behind the choice of this algorithm for this study is twofold. First, XGBoost can handle complex and non-linear relationships among the variables, which we expect to have in a complex phenomenon like food insecurity. Secondly, XGBoost has a high degree of flexibility, which makes it the most suitable candidate for a prediction task that is meant to eventually run as an operational tool in near real-time. Specifically, XGBoost can handle missing values in the input variables, which is a feature that makes it possible to automatically run the algorithm on a daily basis, even when a few values might be missing because, for example, of delays in data availability.

Since XGBoost does not support a multi-output design, we developed 30 different models, one for each prediction horizon. For each date, the prediction framework is trained to predict levels of insufficient food consumption for a given day into the future based on the information available up to the date under consideration. For further details, see Section 6 of the Supplementary Information.

In order to implement our forecasting model based on the usual three stages of training, validation and testing, we adopt a k-fold cross-validation approach in a time-ordered fashion (i.e. the evaluation stage is applied to different historical periods). The validation phase is implemented by splitting each of the *n* splits of training points of each sub-region into two time order preserving sets: the first 80% samples are used for training and the remaining ones for validation. Validation is performed independently across splits ensuring an unbiased approach. Our validation scheme aims to optimize the prediction framework by acting on two main configurations: model hyper-parameters and feature selection. The aim of this optimization is to find the configuration that returns the best performance as measured on a validation set. See Supplementary Table S8 for a detailed list of the explored hyper-parameters and values, and Supplementary Table S7 for the detailed list of independent variables and time lags considered. For further details, see Section 6 of the Supplementary Information.

Finally, in order to assess the goodness of the proposed forecasting model, its performance on the test sets is compared with a naive approach, where the predicted value at any given forecasting horizon is simply given by the last available value in the training and validation set, which represents the last available measured value before the start of the forecasting horizon.

## Data Availability

The data and code used to generate the results reported in this study will be made available at the time of publication.

## Supplementary Information

Supplementary information, figures and tables are available here.

## Disclaimer

The content and views expressed in this paper are solely those of the authors and do not necessarily reflect the official views of the UN World Food Programme.

## Acknowledgements

PF, DP and MT gratefully acknowledge the Lagrange Project of the ISI Foundation funded by the CRT Foundation. EO and GM would like to acknowledge her WFP colleagues for the fruitful discussions.

## Footnotes

* https://dataviz.vam.wfp.org/economic_explorer/prices

† https://dataviz.vam.wfp.org/seasonal_explorer/rainfall_vegetation/help

‡ No investigation into the forecast of the the independent indicators was performed because of the involvement of chain-of-events predictions (e.g. weather or market forecasts) and ethical issues around providing conflict predictions.

§ https://xgboost.readthedocs.io/en/latest/

